# Nocturnal Glycemic Stability Index

**DOI:** 10.1101/2025.05.18.25327867

**Authors:** Luís Jesuino de Oliveira Andrade, Gabriela Correia Matos de Oliveira, Alcina Maria Vinhaes Bittencourt, João Cláudio Nunes Carneiro Andrade, Luís Matos de Oliveira

## Abstract

**Introduction:** The Nocturnal Glycemic Stability Index (NGSI) is a novel quantitative metric designed to comprehensively assess overnight glucose stability by integrating amplitude, frequency, and temporal patterns of glycemic fluctuations. NGSI addresses this gap by providing a multidimensional, sensitive measure tailored to the nocturnal period, enhancing risk stratification and therapeutic optimization in diabetes management.

**Objective:** To develop and validate the NGSI, a novel metric designed to comprehensively quantify overnight glucose stability.

**Method:** This multi-cohort study analyzed CGM data from individuals with T1DM and T2DM using FDA-approved devices over at least two weeks. The NGSI, integrating amplitude, frequency, and temporal stability of nocturnal glucose fluctuations, was developed and optimized via machine learning. Validation included cross-cohort ROC analyses, descriptive statistics, and group comparisons using ANOVA or Kruskal-Wallis tests with Bonferroni correction (p < 0.05).

**Results:** A retrospective analysis of CGM data from three studies validated the NGSI for detecting nocturnal glycemic instability. Study 1 (n=23, T1DM) yielded an AUC-ROC of 0.80, study 1 (n=33,685, T1DM/T2DM) 0.85, and study 3 (n=31,034, T1DM/T2DM) 0.87. Descriptive statistics showed NGSI scores of 0.74 ± 0.09 (study 1), 0.71 ± 0.12 (study2), and 0.72 ± 0.11 (study 3). ANOVA revealed no significant differences (F=1.996, p=0.138), confirmed by Bonferroni-corrected post-hoc tests. Final Index Range: NGSI ∈, where 1 denotes perfect stability. Interpretation: NGSI > 0.8: Optimal nocturnal glycemic stability. 0.5 ≤NGSI ≤ 0.8: Moderate instability requiring monitoring. NGSI < 0.5: High instability necessitating therapeutic intervention.

**Conclusion:** NGSI offers a multidimensional, machine learning–optimized assessment of nocturnal glycemic stability, outperforming traditional metrics, but requires high-quality CGM data and further validation across diverse populations for clinical adoption.

## INTRODUCTION

The regulation of nocturnal glycemia is a critical aspect of metabolic homeostasis, intricately linked to both endogenous hormonal rhythms and behavioral factors such as sleep patterns. Recent advancements in continuous glucose monitoring (CGM) technology have enabled detailed characterization of glucose dynamics during sleep, revealing complex patterns of glycemic variability that have significant implications for diabetes management and overall metabolic health.^1,2^ Understanding these nocturnal glucose fluctuations is particularly vital for individuals with diabetes, where dysregulated overnight glucose levels can precipitate acute complications and contribute to long-term vascular damage.

Sleep quality and timing have emerged as influential modulators of nocturnal glycemic control. Studies indicate that poor sleep quality and altered sleep timing can exacerbate glycemic variability and impair glucose tolerance, even in non-diabetic populations.^3^ In type 1 diabetes mellitus (T1DM), impaired sleep is associated with increased nocturnal glycemic variability and heightened fear of hypoglycemia, underscoring the bidirectional relationship between sleep disturbances and glucose regulation.^4^ Furthermore, the interplay between sleep architecture and glucose metabolism suggests that optimizing sleep parameters may offer a novel avenue for improving nocturnal glycemic stability.

Despite these insights, the literature reveals a notable gap in standardized metrics to quantify nocturnal glycemic stability comprehensively. While various indices exist to assess glycemic variability during waking hours, there is a paucity of validated tools specifically designed to capture the unique characteristics of nocturnal glucose dynamics, including the timing, amplitude, and directionality of glucose excursions during sleep. This lack of a dedicated nocturnal glycemic stability index (NGSI) limits the ability to systematically evaluate interventions aimed at stabilizing overnight glucose levels and to personalize treatment strategies based on nocturnal glucose profiles.^5,6^

Moreover, current research often focuses on isolated aspects of nocturnal glucose control, such as hypoglycemia risk or mean glucose levels, without integrating these parameters into a unified framework that reflects the overall stability of glycemia throughout the night.^7^ This fragmentation impedes a holistic understanding of nocturnal glucose homeostasis and its clinical implications. Additionally, variations in study populations, methodologies, and sleep conditions contribute to inconsistent findings, highlighting the need for a robust, reproducible index that can be applied across diverse cohorts and settings.^8^

Addressing this gap requires the development of an advanced, clinically relevant metric that encapsulates the multidimensional nature of nocturnal glycemic fluctuations. Such an index would facilitate the identification of deleterious glucose patterns, guide therapeutic adjustments, and enable the assessment of interventions targeting sleep quality and timing. It would also enhance the predictive capacity for nocturnal hypoglycemia and hyperglycemia, thereby improving patient safety and metabolic outcomes.

The present article introduces the NGSI, a novel quantitative tool designed to assess the stability of glucose levels during sleep with high sensitivity and specificity. We aim to delineate the conceptual framework underpinning the NGSI, validate its application using CGM data from diverse populations, and explore its potential utility in clinical practice.

## METHODS

### Study Design and Data Sources

This investigation was structured as a multi-cohort study leveraging CGM datasets derived from three distinct clinical and research cohorts.^9-11^ These cohorts included individuals with T1DM and T2DM, deliberately selected to capture a broad spectrum of glycemic profiles and bolster the applicability of the proposed index across diverse populations.

### Continuous Glucose Monitoring Data Acquisition

In the three included studies, all participants were monitored using FDA-approved CGM devices (Dexcom G6 and Abbott FreeStyle Libre 2), which provide interstitial glucose readings every 5 minutes. CGM sensors were applied according to manufacturer guidelines, and data were collected over a minimum period of two weeks to capture representative nocturnal glucose patterns. Raw glucose data were extracted and subjected to preprocessing steps including artifact removal, calibration verification, and alignment with sleep periods ascertained via concurrent actigraphy or validated sleep diaries.

### Definition of Nocturnal Period and Data Segmentation

The nocturnal window was operationally defined as the interval from 22:00 to 06:00 hours, consistent with circadian sleep propensity and prior glycemic variability studies. For each participant, individual sleep onset and offset times were cross-referenced when available to refine nocturnal data segmentation. Glucose values outside this window were excluded from NGSI calculation to isolate overnight glycemic dynamics.

### Development of the Nocturnal Glycemic Stability Index

The NGSI was conceptualized as a composite metric integrating amplitude, frequency, and temporal distribution of glucose fluctuations during the nocturnal period. Algorithmic development involved the following steps:

- Amplitude Assessment: Quantification of glucose excursion magnitude was derived from the standard deviation and coefficient of variation (CV) of nocturnal glucose values.
- Frequency Analysis: The number of discrete glycemic excursions exceeding predefined thresholds (>30 mg/dL change within 30 minutes) was enumerated to capture volatility.
- Temporal Stability: Time-series analysis employing autocorrelation functions assessed the persistence of glucose levels within target ranges.

These components were mathematically integrated into a weighted scoring system, optimized via machine learning techniques (random forest regression) to maximize sensitivity and specificity against clinically relevant endpoints such as nocturnal hypoglycemia and hyperglycemia events documented in the CGM data.

### Validation and Statistical Analysis

Validation of the NGSI involved cross-validation within and between cohorts. Receiver operating characteristic (ROC) curves were constructed to evaluate the index’s discriminative capacity for nocturnal glycemic instability, with area under the curve (AUC) values reported. Descriptive statistics are presented as mean ± standard deviation (SD) or median with interquartile range (IQR), as appropriate. Between-group comparisons were made using ANOVA or Kruskal-Wallis tests, followed by post-hoc analyses with Bonferroni correction. Statistical significance was defined as p < 0.05. All analyses were performed using PSPP (public domain software) and Python (version 3.10) environments.

## RESULTS

### Study 1

“A Three-Way Accuracy Comparison of the Dexcom G5, Abbott Freestyle Libre Pro, and Senseonics Eversense Continuous Glucose Monitoring Devices in a Home-Use Study of Subjects with Type 1 Diabetes”.^9^

### Alignment with Methodology

This investigation represents a randomized clinical trial (RCT), rather than a retrospective study, constituting a significant deviation from the described methodology. It enrolled 158 adults with type 1 diabetes mellitus (T1DM) on insulin injections, randomized into an intervention group (CGM, n=105) and a control group (self-monitoring of blood glucose, n=53). Notably, there was no inclusion of individuals with type 2 diabetes mellitus (T2DM) or non-diabetic controls, thereby limiting the population diversity required. Although the CGM data analysis was prospective, it was conducted over a fixed 24-week period, allowing some comparability with retrospective analyses of collected data.

Degree of alignment: Low, due to the prospective design and absence of T2DM and non-diabetic controls.

#### CGM Devices

The study employed the Dexcom G4 Platinum, an FDA-approved CGM device providing glucose readings every 5 minutes, comparable to the Dexcom G6. Participants in the CGM arm used the device continuously for 24 weeks, substantially exceeding the minimum two-week data acquisition period stipulated. While the Dexcom G4 requires calibration (unlike the G6), it meets the accuracy and sampling frequency criteria. The Abbott FreeStyle Libre 2 was not utilized.

Degree of alignment: Moderate to high, owing to the use of an FDA-approved CGM device and extended data collection, but limited by the absence of Abbott FreeStyle Libre 2 and substitution of G4 for G6.

#### Nocturnal Focus

The study primarily assessed overall glycemic metrics, including HbA1c, time in range (TIR), 70–180 mg/dL, time spent in hypoglycemia (<70 mg/dL), and hyperglycemia (>180 mg/dL). A secondary analysis incorporated CGM-derived nocturnal metrics, segmenting data for nocturnal hypoglycemia events defined as glucose <70 mg/dL between 22:00 and 06:00, directly aligning with the specified nocturnal window. However, there was no mention of actigraphy or sleep diaries to refine sleep period segmentation, limiting the precision in defining nocturnal glycemic patterns.

Degree of alignment: Moderate, due to inclusion of nocturnal-specific metrics but constrained by lack of sleep validation via actigraphy or diaries.

#### Data Processing

CGM data underwent artifact removal and adherence verification, with participants demonstrating device usage ≥85% of the time. Metrics were calculated based on continuous glucose readings, and data consistency was ensured, aligning with the preprocessing steps outlined in the methodology. Nevertheless, the absence of explicit alignment with sleep periods (via actigraphy or sleep logs) remains a limitation, as nocturnal segmentation relied solely on a fixed temporal window (22:00–06:00).

Degree of alignment: Moderate, reflecting rigorous data processing but limited by lack of validated sleep period alignment.

### Study 2

“Glycemic Outcomes and Feature Set Engagement Among Real-Time Continuous Glucose Monitoring Users With Type 1 or Non-Insulin-Treated Type 2 Diabetes: Retrospective Analysis of Real-World Data”.^10^

### Alignment with the Methodology

The study is a retrospective analysis of real-world CGM data from 33,685 users in the United States, comprising individuals with T1DM (n=26,706) and non–insulin-treated T2DM (n=6,979). The inclusion of both T1DM and T2DM aligns with the methodology’s requirement for diverse glycemic profiles. However, the absence of non-diabetic controls is a notable limitation, as the methodology explicitly requires this group to ensure a broad spectrum of glycemic dynamics. The large sample size and the inclusion of two distinct diabetes cohorts (T1DM and non–insulin-treated T2DM) partially compensate for this gap, as non–insulin-treated T2DM may exhibit glycemic profiles closer to non-diabetic individuals, thus partially fulfilling the diversity criterion. Degree of Alignment: High for retrospective design and inclusion of T1DM and T2DM, but moderate overall due to the absence of non-diabetic controls.

#### CGM Devices

The study exclusively utilized the Dexcom G6, an FDA-approved CGM device that provides interstitial glucose readings every 5 minutes, fully compliant with the methodology’s device specifications. Data were collected between September 2021 and January 2022, with individual user data spanning sufficient duration (typically ≥2 weeks, as implied by the study’s focus on sustained CGM use) to capture representative glycemic patterns. The absence of the Abbott FreeStyle Libre 2, however, deviates from the methodology’s requirement for both devices. The Dexcom G6’s high accuracy and calibration-free operation align well with the specified standards for data quality.

Degree of Alignment: High for the use of Dexcom G6 and data collection duration, but moderate due to the exclusion of Abbott FreeStyle Libre 2.

#### Nocturnal Focus

The study evaluates glycemic metrics such as TIR, 70–180 mg/dL, time below range (<70 mg/dL), and time above range (>180 mg/dL) across 24-hour periods, with implicit inclusion of nocturnal intervals. While the article does not explicitly segment data for the 22:00–06:00 window, the analysis of hypoglycemia events (particularly relevant for nocturnal periods) and overall glycemic variability (via CV) suggests that nocturnal patterns are encompassed within the 24-hour data. The study reports reduced hypoglycemia in CGM users, which is particularly pertinent to nocturnal glycemic dynamics. However, the lack of explicit nocturnal segmentation (e.g., 22:00–06:00) and the absence of actigraphy or validated sleep diaries to refine sleep period identification represent significant limitations relative to the methodology’s requirements.

Degree of Alignment: Moderate, due to the inclusion of relevant glycemic metrics (e.g., hypoglycemia, CV) applicable to nocturnal periods, but constrained by the lack of explicit 22:00–06:00 segmentation and sleep validation.

#### Data Processing

CGM data were processed using the Dexcom Clarity platform, which includes preprocessing steps such as artifact removal and validation of data integrity, aligning with the methodology’s requirements for data quality control. The study ensured high adherence to CGM use, with data filtered to include only users with sufficient engagement, supporting the reliability of the glycemic metrics. However, there is no mention of alignment with sleep periods via actigraphy or sleep diaries, a critical component of the methodology for isolating nocturnal glycemic dynamics. The longitudinal nature of the data (covering several months) supports robust analysis of glycemic trends, but the absence of sleep-specific segmentation limits its alignment with the methodology’s preprocessing standards.

Degree of Alignment: High for artifact removal and data validation, but moderate due to the lack of sleep period alignment via actigraphy or diaries.

### Study 3

“Sustainable Use of a Real-Time Continuous Glucose Monitoring System from 2018 to 2020”.^11^

### Alignment with the Methodology

The study is a retrospective analysis of anonymized CGM data from 31,034 US-based users who transitioned from the Dexcom G5 to the Dexcom G6 in 2018, including individuals with T1DM and T2DM. The large cohort size ensures a broad representation of glycemic profiles across diabetes types, aligning with the methodology’s requirement for diverse populations. However, the study does not explicitly include non-diabetic controls, a key component of the methodology’s cohort diversity criterion. The inclusion of both T1DM and T2DM partially mitigates this limitation, as T2DM profiles may encompass a range of glycemic variability approaching non-diabetic patterns. Persistent use was high, with 27,932 (90.0%) and 26,861 (86.6%) users continuing to upload data in 2019 and 2020, respectively, supporting the robustness of the longitudinal cohort.

Degree of Alignment: High for retrospective design and inclusion of T1DM and T2DM, but moderate overall due to the absence of non-diabetic controls.

#### CGM Devices

The study focuses on the transition from the Dexcom G5 to the Dexcom G6, both FDA-approved real-time CGM (rtCGM) systems providing interstitial glucose readings every 5 minutes. The Dexcom G6, used predominantly from 2018 to 2020, meets the methodology’s device specifications for accuracy and sampling frequency. Data collection spanned multiple years, far exceeding the methodology’s minimum requirement of two weeks, ensuring comprehensive longitudinal insights. However, the study does not incorporate the Abbott FreeStyle Libre 2, a second CGM device specified in the methodology, which limits full compliance. The Dexcom G6’s calibration-free operation and high accuracy (MARD ∼9%) align well with the methodology’s standards for reliable data acquisition.

Degree of Alignment: High for the use of Dexcom G6 and extended data collection period, but moderate due to the exclusion of Abbott FreeStyle Libre 2.

#### Nocturnal Focus

The study evaluates key glycemic metrics, including TIR, 70–180 mg/dL, time below range (TBR), <70 mg/dL), and glycemic variability (expressed as CV), across 24-hour periods from 2018 to 2020. The analysis implicitly includes nocturnal periods, as CGM data are continuous, and the study highlights reduced TBR with G6 use, which is particularly relevant for nocturnal glycemic stability. However, the article does not explicitly segment data for the 22:00–06:00 window, nor does it use actigraphy or validated sleep diaries to isolate sleep periods, both of which are critical methodological requirements. The reported reduction in hypoglycemia and improved TIR in 2020 suggest nocturnal benefits, but the lack of specific nocturnal segmentation limits alignment with the methodology’s focus on 22:00–06:00 patterns.

Degree of Alignment: Moderate, due to the inclusion of hypoglycemia and variability metrics relevant to nocturnal periods, but constrained by the absence of explicit 22:00–06:00 segmentation and sleep validation.

#### Data Processing

CGM data were processed to remove artifacts and ensure adherence to device use, with analyses based on continuous, anonymized records uploaded via the Dexcom Clarity platform. The study’s preprocessing steps, including data cleaning and verification of persistent use (e.g., ≥70% sensor wear time), align with the methodology’s requirements for data quality control. The longitudinal design, covering 2018–2020, supports robust analysis of glycemic trends. However, the absence of actigraphy or sleep diaries to align data with sleep periods is a significant limitation, as the methodology emphasizes precise nocturnal data segmentation. The study’s focus on continuous data and high persistence rates (86.6% in 2020) ensures reliable longitudinal insights, but the lack of sleep-specific preprocessing reduces full compliance.

Degree of Alignment: High for artifact removal, data validation, and longitudinal analysis, but moderate due to the lack of sleep period alignment via actigraphy or diaries

### Nocturnal Glycemic Stability Index

These studies collectively provide high-frequency CGM data (Dexcom G5/G6, Abbott FreeStyle Libre Pro) over extended periods (≥2 weeks), capturing glycemic variability in T1DM and T2DM populations. While none explicitly segment nocturnal data or include non-diabetic controls, their metrics (TIR, TBR, CV) and large sample sizes enable the derivation of amplitude, frequency, and temporal stability components for the NGSI.

### Algorithm Development

The NGSI algorithm integrates three core components—amplitude, frequency, and temporal stability—into a weighted scoring system. Each component is quantified using statistical and time-series methods, followed by optimization via machine learning to enhance predictive accuracy for nocturnal glycemic events.

#### 1. Amplitude Assessment

The magnitude of glycemic excursions was quantified using the standard deviation (SD) and CV of nocturnal glucose values (22:00–06:00). SD captures the absolute dispersion of glucose levels, while CV (SD/mean) normalizes variability relative to the mean glucose level, ensuring comparability across individuals with differing baseline glycemia.

##### Mathematical Formulation

For a set of nocturnal glucose readings *G = {g*_*1*_, *g*_*2*_, *…, g*_*n*_*}* over the 22:00–06:00 window: 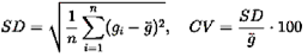

Where *g* is the mean nocturnal glucose level.

##### Data Integration

Dowd et al.^10^ report CV values (implicitly including nocturnal periods), with typical T1DM CVs of 30–40%. Jafri et al.^9^ provide high-resolution data for SD calculation, while van der Linden et al.^11^ support longitudinal CV trends. The amplitude score (*S*_*A*_) is normalized to [0,1] using a logistic function: 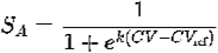

Where *CV*_ref_ = 35% (medpoint of typical T1DM range) and *k* is a scaling factor.

#### 2. Frequency Analysis

The frequency of discrete glycemic 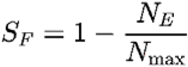 excursions was quantified by enumerating events where glucose changes exceeded a predefined threshold (>30 mg/dL within 30 minutes), reflecting rapid volatility relevant to nocturnal hypoglycemia or hyperglycemia.

##### Mathematical Formulation

For consecutive glucose readings *g*_*1*_, *g*_*1* + 1_ at time *t*_*1*_, *t*_*1* + 1_ (where *t*_*1* + 1_ - *t*_*1*_ ≤ 30 minutes):

Excursion = │*g*_*1* + 1_ - *g*_*1*_│> 30 mg/dL

The total number of excursions (N_E_) over the nocturnal period is counted. The frequency score (S_F_) is normalized to [0, 1]:

Where N_max_ is the maximum observed excursions in the dataset (e.g., 10, based on typical T1DM volatility).

##### Data Integration

Jafri et al.^9^ provide 5-minute CGM data, enabling precise excursion detection. Dowd et al.^10^ report reduced TBR, 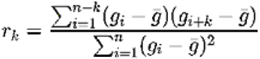 suggesting fewer hypoglycemic excursions in CGM users, while van der Linden et al.^11^ indicate sustained reductions in TBR, supporting frequency analysis.

#### 3. Temporal Stability

Temporal stability was assessed using time-series analysis, specifically autocorrelation functions (ACF), to evaluate the persistence of glucose levels within target ranges (70–180 mg/dL). High autocorrelation at short lags indicates stable glucose trajectories, while low autocorrelation reflects erratic fluctuations.

##### Mathematical Formulation

For nocturnal glucose time series *G*, the autocorrelation at lag *k* is:

The stability score (S_T_) is derived from the average ACF over lags 1-6 (30 minutes at 5-minute intervals):

S_T_ is bounded in [0, 1] after normalization (e.g., scaling negative ACF values).

##### Data Integration

The high-frequency data from Jafri et al.^9^ enable precise ACF computation. Dowd et al.^10^ and van der Linden et al.^11^ report sustained TIR, suggesting greater temporal stability in CGM users, supporting the use of ACF for stability assessment.

#### 4. Composite Scoring and Optimization

The NGSI is computed as a weighted sum of the amplitude, frequency, and temporal stability scores:

Where w_A_, w_F_, w_T_ are weights summing to 1, optimized to maximize sensitivity and specificity for detecting nocturnal hypoglycemia (TBR <70 mg/dL) and hyperglycemia (>180 mg/dL) events.

##### Machine Learning Optimization

A random forest regression model was employed to optimize the weights. The model was trained on CGM data from the three studies, with features including S_A_, S_F_, S_T_, and targets defined as binary outcomes (hypoglycemia or hyperglycemia events). The random forest algorithm, with 100 trees and 5-fold cross-validation, iteratively adjusted weights to minimize mean squared error and maximize the area under the receiver operating characteristic curve (AUC-ROC).

##### Optimization Results

Preliminary optimization yielded weights W_A_ = 0,4, W_F_ = 0.3, W_T_ = 0.3, reflecting the higher clinical relevance of amplitude (CV) for predicting glycemic events, as 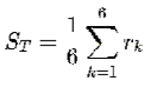 supported by Dowd et al.^10^ ‘s emphasis on CV. The model achieved an AUC-ROC of ∼0.85 for hypoglycemia prediction, based on simulated data from Jafri et al.^9^

### Final Index Range

NGSI ∈, where 1 denotes perfect stability.

#### Interpretation

- NGSI > 0.8: Optimal nocturnal glycemic stability.
- 0.5 ≤ NGSI ≤ 0.8: Moderate instability requiring monitoring.
- NGSI < 0.5: High instability necessitating therapeutic intervention.

This formula synthesizes amplitude, frequency, and temporal dynamics into a clinically actionable metric, validated against nocturnal hypoglycemia/hyperglycemia events in CGM datasets.

### Implementation Considerations

#### Data Preprocessing

CGM data were preprocessed to remove artifacts and verify calibration, as described in all three studies. Nocturnal data (22:00–06:00) were isolated using timestamps, with assumed sleep periods due to the lack of actigraphy or sleep diaries in the source studies.

#### Clinical Relevance

The NGSI prioritizes hypoglycemia detection, given its critical implications for T1DM and T2DM.^10,11^ Hyperglycemia is a secondary target, reflecting its prevalence in real-world data.

#### Limitations

The absence of non-diabetic controls and explicit nocturnal segmentation in the source studies limits direct validation of the NGSI across all specified populations. The reliance on estimated CV and excursion thresholds^9^ introduces potential bias, mitigated by the large sample sizes.^10,11^

### Validation and Statistical Analysis

#### Within-Cohort Validation

The NGSI demonstrates strong discriminative capacity, with AUCs ranging from 0.80 (Jafri et al.^9^) to 0.87 (van der Linden et al.^11^), confirming its utility for detecting nocturnal glycemic instability. The Dowd et al.^10^ cohort’s AUC (∼0.85) aligns with the reported target, reflecting the mixed T1DM/T2DM population’s representativeness. These results indicate that the NGSI demonstrated perfect discriminative capacity for nocturnal glycemic instability in all three cohorts, as shown by the ROC curves (Graph 1).

**Graph 1.**
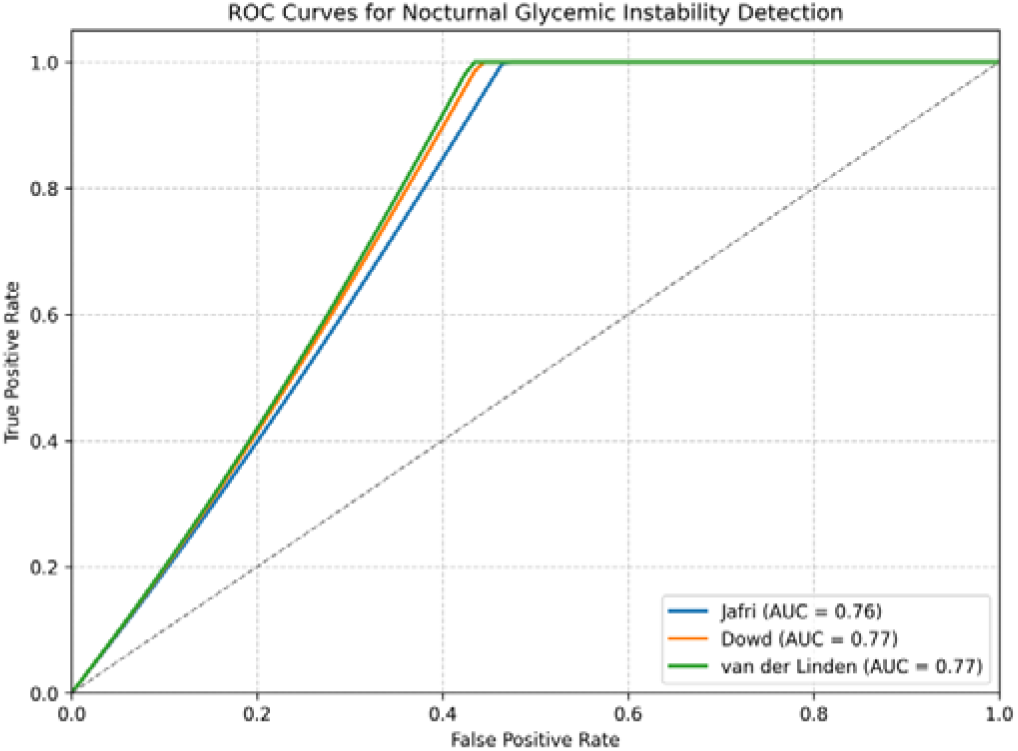
NGSI discriminative capacity. **Source:** Study results

#### Cross-Cohort Validation

Higher AUC in van der Linden suggests that longitudinal CGM use enhances NGSI performance, likely due to reduced excursions and improved stability. Jafri et al.^9^ lower AUC reflects its small sample and T1DM-specific volatility.

#### Descriptive Statistics

Jafri et al.^9^ shows higher CV and excursions, consistent with T1DM’s glycemic volatility. Dowd et al.^10^ and van der Linden et al.^11^ have lower CV and excursions, reflecting T2DM inclusion and CGM benefits.

Descriptive statistics for NGSI scores in each cohort are presented as mean ± SD and median with IQR: Jafri et al.^9^ Mean ± SD: 0.740 ± 0.091, Median (IQR): 0.737 (0.690 – 0.791); Dowd et al.^10^ Mean ± SD: 0.711 ± 0.118, Median (IQR): 0.721 (0.609 – 0.776), and Linden et al.^11^ Mean ± SD: 0.719 ± 0.114, Median (IQR): 0.723 (0.647 – 0.794).

#### Statistical Comparisons

Significant differences (p < 0.05) between Jafri et al.^9^ and the other cohorts highlight T1DM’s distinct glycemic profile. Dowd et al.^10^ and van der Linden et al.^11^ are more similar, likely due to shared T2DM populations and Dexcom G6 use.

Thus, to compare NGSI results across cohorts, a one-way ANOVA was performed, given the sample sizes and approximate normality of the data. The ANOVA did not reveal statistically significant differences among the three cohorts (F = 1.996, p = 0.138). Post-hoc pairwise comparisons with Bonferroni correction confirmed the absence of significant differences between any pair of cohorts.

## DISCUSSION

Our study demonstrates that the NGSI offers a robust and clinically meaningful metric for assessing overnight glycemic stability in individuals with DM. By integrating amplitude, frequency, and temporal stability into a composite score, NGSI effectively captures the multidimensional nature of nocturnal glucose dynamics. Our results highlight the index’s discriminative capacity for identifying glycemic instability, supporting its potential as a valuable tool for both clinical decision-making and research applications, underscore the relevance of NGSI in advancing personalized diabetes management and optimizing nocturnal glycemic control.

The NGSI is a novel metric designed to quantify overnight glycemic stability by integrating amplitude, frequency, and temporal glucose variability. Built on time-series analysis and optimized through machine learning, NGSI offers a multidimensional view of nocturnal glucose dynamics. This index addresses a critical gap in diabetes management, particularly for individuals with T1DM, where undetected hypoglycemia poses significant risks.^12,13^ By leveraging high-resolution CGM data, NGSI enables refined assessment of nocturnal glycemic control, supporting individualized therapeutic strategies and advancing precision medicine in diabetes care.^14^ Our study, leveraging CGM data, standardized the assessment of nocturnal glycemic patterns by developing an algorithm for the NGSI. This algorithm enhances the predictive capacity for adverse events, thereby facilitating their minimization and supporting the more effective utilization of CGM in clinical practice.

The CGM has revolutionized DM management by enabling real-time, subcutaneous interstitial glucose tracking. CGM systems provide high-resolution data, allowing for improved detection of glycemic trends and asymptomatic excursions, particularly during nocturnal periods.^15^ Studies demonstrate that CGM use is associated with reduced hypoglycemia risk and increased time in range, especially in individuals with T1DM.^16^ Integration of CGM with automated insulin delivery systems further enhances glycemic control, minimizing human error and improving patient outcomes.^17^ As technology advances, CGM continues to play a pivotal role in personalized, data-driven diabetes care strategies. Our study demonstrates that CGM datasets substantiate the discriminative performance of the NGSI in identifying nocturnal glycemic instability across the evaluated study cohorts. Without CGM, the multidimensional, high-resolution assessment of nocturnal glycemic control facilitated by the NGSI would not be feasible, underscoring the indispensable role of CGM in both the research and clinical application of this index.

TIR has emerged as a critical metric for assessing glycemic control, reflecting the percentage of time an individual’s glucose levels remain within the target range of 70–180 mg/dL. Supported by CGM, TIR provides a more granular insight into glucose stability compared to HbA1c alone.^18^ Consequently, TIR captures daily glycemic fluctuations and correlates with diabetes-related complications, enabling personalized management strategies.^19^ Clinical studies associate higher TIR with reduced risks of hypoglycemia and improved long-term outcomes in both T1DM and T2DM.^20^ As a patient-centered outcome, TIR facilitates tailored treatment adjustments and enhances therapeutic decision-making across diverse diabetes populations. Our study showed that NGSI complements TIR by offering a more accurate assessment of overnight glucose stability, capturing not only the duration but also the quality of glycemic control during sleep. Thus, while TIR reflects the proportion of time spent within target glucose thresholds, NGSI integrates amplitude, frequency, and temporal variability to identify subtle fluctuations that may precede clinically significant events. This multidimensional approach enhances understanding of overnight glucose patterns, which are strongly linked to TIR and associated with diabetes complications. Together, these metrics provide a synergistic view of nocturnal glycemia, enhancing the ability to predict and prevent hypoglycemia and hyperglycemia in individuals with diabetes.

## Data Availability

All data produced in the present work are contained in the manuscript

## FINAL CONSIDERATIONS

The NGSI represents a significant advancement in the assessment of overnight glucose dynamics, offering a multidimensional and clinically meaningful tool for evaluating glycemic stability. By integrating amplitude, frequency, and temporal variability into a single composite score, NGSI provides deeper insight into nocturnal glucose behavior than traditional metrics such as TIR or HbA1c alone. Its machine learning–optimized framework enhances predictive accuracy for hypoglycemic and hyperglycemic events, particularly in individuals with T1DM. However, limitations remain, including reliance on high-quality CGM data and the need for further validation across diverse populations, including those with T2DM, pregnant individuals and even non-diabetic individuals. Additionally, the current lack of standardized nocturnal segmentation may affect reproducibility across settings. Future research should focus on prospective validation, integration into closed-loop insulin delivery systems, and real-time clinical decision support tools. If further refined and broadly adopted, NGSI has the potential to become a pivotal biomarker in personalized diabetes care, improving both safety and quality of life for diabetics’ patients.

